# COVID -19: could green tea catechins reduce the risks?

**DOI:** 10.1101/2020.10.23.20218479

**Authors:** Maksim Storozhuk

**Author notes:** Corresponding author Dr. M. Storozhuk, Bogomoletz Institute of Physiology, National Academy of Science of Ukraine, 4 Bogomoletz Street, Kiev 01024 Ukraine.

## Abstract

**Purpose:** Several lines of emerging pharmacological and epidemiological evidence imply that overall risks related to COVID-19 may be reduced by green tea catechins. Therefore, it may be expected that countries with higher per/capita green tea consumption would be less affected by COVID-19. The aim of this study was to assess this possibility.

**Methods:** Among countries with at least 3 million population (n=134), countries with relatively high (above 150 g) per/capita green tea consumption have been identified (n=21); (ii) normalized to population values of COVID-19 cases (morbidity) and deaths (mortality) for groups of countries with high and low per/capita green tea consumption were compared.

**Results:** Striking differences in COVID-19 morbidity and mortality between groups of countries with ‘high’ and ‘low’ green tea consumption were found. The differences were still observed after the adjustment for the onset of the disease. An analysis using multiple linear regression approach suggests that the associations are present at the level of individual countries.

**Conclusion:** Evidence supporting the idea that green tea constituents could reduce overall risks related to COVID-19 has been obtained. The results are promising and are in line with emerging evidence from other studies including pharmacological ones. Nevertheless, because of limitations of this study the idea still should be considered as a hypothesis requiring further assessment. Several vaccines are currently validated for COVID-19 prevention and mass vaccination has already been started in many countries. Still, it is likely that the development of an efficient drug therapy that reduces COVID-19 severity/mortality would be important for rather prolonged time. In this context, the results obtained in this study may have significant implications.

## Introduction

Several vaccines are currently validated for COVID-19 prevention and mass vaccination has already been started in some countries. Nevertheless, it is likely that development of efficient drug therapy that reduces COVID-19 severity/mortality would be important for rather prolonged time. Several lines of evidence suggest that green tea catechins (such as epigallocatechin-3-gallate (EGCG)) could give a clue to this issue. As briefly outlined below this could be due to (i) direct anti-viral activity and (ii) the influence on factors associated with COVID-19 severity and mortality.

Green tea catechins, are known to have anti-viral activity against several viruses [1-4] including viruses causing respiratory diseases [5, 6]. Recent target-based virtual ligand screening study suggests that at least one catechin (EGCG), is likely to target papain-like proteinase (PLpro), an indispensable enzyme in the process of coronavirus replication and infection of the host [7]. Inhibition of SARS-CoV-2 3CL-Protease by EGCG has been recently demonstrated in *vitro* [8]. Additionally, several factors associated with COVID-19 mortality are likely to be affected by green tea constituents. Indeed, there is evidence that green tea catechins: lower cholesterol levels [9, 10]; have anti-diabetic [11] and anti-obesity effects [12]; are beneficial in cardiovascular disease [13].

Besides, EGCG is protective in diseases with uncontrolled immune activation, thus, may ameliorate severity of COVID-19 [14] since the latter is related to overreaction of the immune system [15]. Finally, green tea catechins can act as ionophores for zinc ions, while the latter are considered as potentially beneficial in relation to COVID-19 [16, 17].

Altogether these results suggest that green tea catechins could reduce overall risks related to COVID-19. If so, it may be expected that countries with higher level of green tea consumption are less affected by COVID-19.

To address this hypothesis an ecological analysis was performed in this study. For this purpose: (i) among countries with at least 3 million population (n=134), countries with relatively high (above 150 g) per/capita green tea consumption have been identified (n=21); (ii) normalized to population values of COVID-19 cases (morbidity) and deaths (mortality) for groups of countries with high and low per/capita green tea consumption were compared.

## Methods

### Ethics approval and consent to participate

Not applicable (ethical approval was not deemed necessary as this was an analysis of publicly available data).

### Identification of countries with relatively high per/capita green tea consumption

It has been previously reported that higher per/capita green tea consumption is typical for some countries in Asia, North Africa and Middle East [18]. Values for several countries in Asia (China, Vietnam, Japan and Indonesia) have been reported [19]. There is also information regarding Taiwan, arbitrary considered as a green tea consuming in the framework of this work because of predominant consumption of the Paochong tea (oxidized by only (8-12%) and Oolong tea (semioxidised)[20]. The value of green tea consumption in Taiwan, indicated in the Table 1 (750 g) was arbitrary estimated as equal to one half of that reported [21] for *total* tea consumption in this country. The relevant and reliable information regarding green tea consumption in the other countries is scarce [22], it is not as trivial as it may seem to find it. Therefore, to estimate the values of per/capita green tea consumption in the other countries the following approach was used. It was assumed that the difference between annual green tea import and export in a particular country (except countries with considerable green tea production) is roughly equivalent to annual green tea consumption. Thus, the difference between import and export divided by population should give a reasonable estimate of per/capita consumption. Indeed, such an approach is used for estimation for countries that do not produce tea [22].

**Table 1.** A tentative list of countries with ‘high’ (above 150 g per/capita) green tea consumption.

|  | Country | Per capita green tea consumption (g) | COVID-19 morbidity (per million population) | COVID-19 mortality (per million population) |
| --- | --- | --- | --- | --- |
| 1 | Mauritania | 3393 | 3386 | 85 |
| 2 | Morocco | 1783 | 12319 | 212 |
| 3 | Taiwan | 750 | 36 | 0.3 |
| 4 | Afghanistan | 719 | 1370 | 59 |
| 5 | Japan | 650 | 2449 | 34 |
| 6 | Mali | 605 | 379 | 15 |
| 7 | Mongolia | 552 | 457 | 0.6 |
| 8 | Uzbekistan | 525 | 2310 | 18 |
| 9 | Libya | 451 | 15606 | 239 |
| 10 | Vietnam | 380 | 16 | 0.4 |
| 11 | Senegal | 345 | 1319 | 30 |
| 12 | Algeria | 332 | 2335 | 64 |
| 13 | China | 320 | 61 | 3 |
| 14 | Hong Kong | 290 | 1262 | 22 |
| 15 | Tunisia | 247 | 14729 | 465 |
| 16 | Guinea | 219 | 1056 | 6 |
| 17 | Niger | 212 | 166 | 6 |
| 18 | Burkina Faso | 167 | 419 | 5 |
| 19 | Indonesia | 200 | 3259 | 94 |
| 20 | UAE | 196 | 25094 | 74 |
| 21 | Denmark | 178 | 32278 | 296 |
| World<sup>1</sup> (average) |  |  | 18378 | 314 |

Per capita green tea consumption is a coarse estimate (see Methods for details). In relation to further analysis, information regarding COVID-19 morbidity and mortality is also provided. This information was retrieved from ‘Worldometer’ [24] as provided on January 16, 2021. World1 – refers to top 212 countries and territories (ranked by population) affected by COVID-19 according to information from ‘Worldometer’ [24] as provided on January 16, 2021.

The information about green tea import and export for particular countries was obtained from Commodity Trade Statistics Database | United Nations Statistics Division database [23], the search was limited to records since the year 2010. Since recent records are unavailable for some countries, to minimize time bias, it was arbitrary chosen to use values for year 2016 as a standard (still for a few countries older records in the database were used in this work).

Using reported values for China, Vietnam, Japan and Indonesia [19], an estimate for Taiwan and the approach mentioned above it has been found that per/capita green tea consumption is above 150 grams in twenty one countries. However, it is essential to note that the list (Table 1) is probably incomplete (see considerations below), because the approach used for the estimate has several limitations. Namely, it cannot be excluded that some countries with considerable green tea per/capita consumption have not been identified among those producing green tea (for instance, Myanmar, South Korea, Papua New Guinea). Additionally, some countries may have not been identified because of absence of the relevant records in the UN database (for instance, Tajikistan). Finally, the estimates may be biased by import/export operations not reflected in the UN database. This is especially likely for ‘smaller’ import/export operations and thus for less populated countries. For this reason, detailed screening of the database was limited to countries with at least a 3 million population.

### Data regarding COVID-19 morbidity and mortality

All data were obtained from open resources. Information on total number of cases and total number of deaths was obtained from ‘Worldometers info. Coronavirus’[24]. The information on Worldometer is based on official daily reports and considered as a reliable (for instance, [25] [26]. For some comparisons, information about COVID-19 morbidity (defined as number of cases per million population) and mortality (defined as number of deaths per million population) for a specific date was directly obtained from [24]. For others, considering that the onset of the disease was not simultaneous in different countries, an adjustment for this factor was made as described below. Date when the total number of confirmed cases in a particular country became 50 (or more) was determined based on information provided in [24] and considered as a reference time point (referred further as the onset date). Then information about the total number of cases/deaths at a date eight months after the onset date was obtained from [24] and normalized to population. These adjusted values were (also) used for comparisons. Eight-month period after the onset date has been chosen as a compromise between longer period for analysis and a number of countries available for analysis (performed in January 2021). Since in some countries the onset occurred in June and even July (2020) some countries were excluded from a comparison of the *adjusted* values. Calculated values reported in the tables are (typically) rounded to an integer, while values used for statistical comparisons were not rounded.

### Statistical analysis

Since the variables of COVID-19 morbidity and COVID-19 mortality do not have a normal distribution [25], non–parametric statistic was primarily employed for the analysis (as suggested in [25, 27]). Namely, Wilcoxon (Mann-Whitney U Test) for Unpaired Data and Spearman Rank Correlation were used for the analysis. On the hand, for multiple linear regression analysis, an approach similar to that previously reported [25] was used. In this analysis, morbidity and mortality per million of population were transformed to the common logarithm (log10) to adjust for normality of the distribution as suggested previously [25].

Factors, included in multiple linear regression analysis (beside green tea consumption) were: population density [28], percentage of population aged above 65 [29], percentage of urban population [30]. In a complementary analysis an additional variable, namely Human Developmental Index (HDI) based on access to health and education services and income [31] was added to the model. ‘KyPlot’ software was employed for statistical assessments.

## Results

A tentative list of countries with relatively high (above 150 g) per/capita green tea consumption was created using the approach described in Methods by screening relevant information for countries and territories listed in [24] *and* population with at least three million (134 countries and territories). Among those 21 were identified as countries with ‘high’ (above 150 g) per/capita green tea consumption (**Table 1**).

Among the other 113 countries: in 18 the overall annual balance between import and export of green tea was negative; for 13 countries no records were found in the UN database (since year 2010). Since information about *production* of green tea in countries with negative import/balance was not available, the group of these 113 countries was considered as a group with (i) low and (ii) undetermined consumption. Finally, a group of countries with the *estimated* consumption below 150 g was considered as a ‘low-consuming’ (n=82). Information about total number of COVID-19 cases (per one million population) and total number of COVID-19 death (per one million population) was obtained from ‘Worldometer’ [24] as provided on January 16, 2021 for the countries from the above mentioned three groups and summarized in **Table 2**.

**Table 2.** COVID-19 morbidity and mortality in relation to per/capita green tea consumption.

|  | Group 1<br>(countries with ‘high’<br>green tea consumption)<br>N=21 | Group 2<br>(countries with ‘low’ or<br>undetermined green tea<br>consumption)<br>N=113 | Group 3<br>(countries with<br>‘low’ green tea<br>consumption)<br>N=82 | World<sup>1</sup> |
| --- | --- | --- | --- | --- |
| COVID-19<br>morbidity | 1370<br>(419-3386) | 10056<sup>**</sup><br>(930-35731) | 15601<sup>**</sup><br>(1213-36434) | 8736 |
| COVID-19<br>mortality | 30<br>(6-85) | 166<sup>**</sup><br>(15-732) | 286<sup>***</sup><br>(26-868) | 94 |

Values (per one million of population) are: median and interquartile range (IQR). \*\* (P<0.01) and \*\*\* (P<0.001) denote significance level of difference as compared to Group 1 (Wilcoxon (Mann-Whitney U Test) for Unpaired Data). Raw data for individual countries are from ‘Worldometer’ [24] as provided on January 16, 2021. World1 – refers to top 212 countries and territories (ranked by population) affected by COVID-19 according to information from ‘Worldometer’ [24] as provided on January 16, 2021.

Both, COVID-19 morbidity and mortality were strikingly (several times) lower in the Group 1 as compared to Group 2 (and Group3). Besides, the differences were highly significant (**Table 2**). These results support the hypothesis that green tea constituents could reduce overall risks related to COVID-19. On the other hand, this preliminary analysis has several limitations. In particular, the onset of the disease was not simultaneous in different countries. To adjust for this the approach described in Method section was used.

A summary for the adjusted values is shown in **Table 3**. Notice a reduction of ‘N’ in the groups as compared to Table 2. This is due to late onset of the disease in some of the countries (values for four countries in Group 2 and two countries in Group 3 were not available at the time of analysis (January 22, 2021)). There were still profound and statistically significant differences in COVID-19 morbidity and mortality between Group 1 and Group 2/Group 3 (**Table 3**).

**Table 3.** Adjusted for the onset COVID-19 morbidity and mortality in relation to per/capita green tea consumption.

|  | Group 1<br>(countries with ‘high’ green<br>tea consumption) | Group 2<br>(countries with ‘low’ or<br>undetermined green tea<br>consumption) | Group 3<br>(countries with ‘low’ green<br>tea consumption) |
| --- | --- | --- | --- |
|  | N=21 | N=109 | N=80 |
| COVID-19 | 996 | 6767** | 10054*** |
| morbidity | (260-3433) | (894-15891) | (1077-17640) |
| COVID-19 | 18 | 109** | 155*** |
| mortality | (3-55) | (15-307) | (24-387) |

Values (per one million of population) are: median, IQR. \*\* (P<0.01) and \*\*\* (P<0.001) denote significance level of difference as compared to Group 1 (Wilcoxon (Mann-Whitney U Test) for Unpaired Data). Values represent the epidemic situation eight months after the onset of the disease (see Methods for more details) for the countries with known values as of January 22, 2021.

To check for possible association between per capita green tea consumption and COVID-19 morbidity/mortality at the level of individual countries, corresponding correlation coefficients were calculated. Namely, Spearman Rank Correlation was used to calculate correlation coefficients between: (i) the *adjusted* for the onset value of morbidity (or mortality) and (ii) per capita green tea consumption in a particular country. Thus, the total number of countries considered in this analysis was 101 (see Supplementary **Table S1)**. No statistically significant correlation between green tea consumption and the morbidity was observed (Rho=-0.12; P=0.22), while there was a weak but significant correlation between estimated green tea consumption and the mortality (Rho=-0.2; P=0.04).

To further study potential associations a linear regression analysis accounting for several potential confounders was used. In this analysis: (i) morbidity and mortality per million of population were transformed to the common logarithm (log10) to adjust for normality of the distribution (as suggested in [25]); (ii) in addition to green tea consumption, factors, reported previously as important confounders were included. In regard of the latter: population density [32]; percentage of population aged above 65 [32] and percentage of urban population [25] were included. Mauritania was excluded from the linear regression analysis as the apparent outlier in terms of estimated green tea consumption. Indeed, value of 3393 g/capita is much greater than Q3+1.5*IQR for the *group with high green tea consumption* (where Q3 is 3-rd quartile, IQR interquartile range), thus was considered as outlier according to Tukey’s rule [33]. Additionally, since the number of deaths (per million) in Cambodia was 0 (as of January 22, 2021) and could not be transformed to the logarithm, this country was excluded from the analysis in regard of the *mortality*.

It has been found that there are: weak but statistically significant correlations between morbidity (and mortality) and green tea consumption; stronger correlations between morbidity (and mortality) and percentage of urban population. Plots for COVID-19 morbidity and mortality in relation to these two factors are shown in **Fig. 1**, additional details about results of the analysis are provided in Supplementary **Table S2 (A**,**B)**.

**Figure 1.**
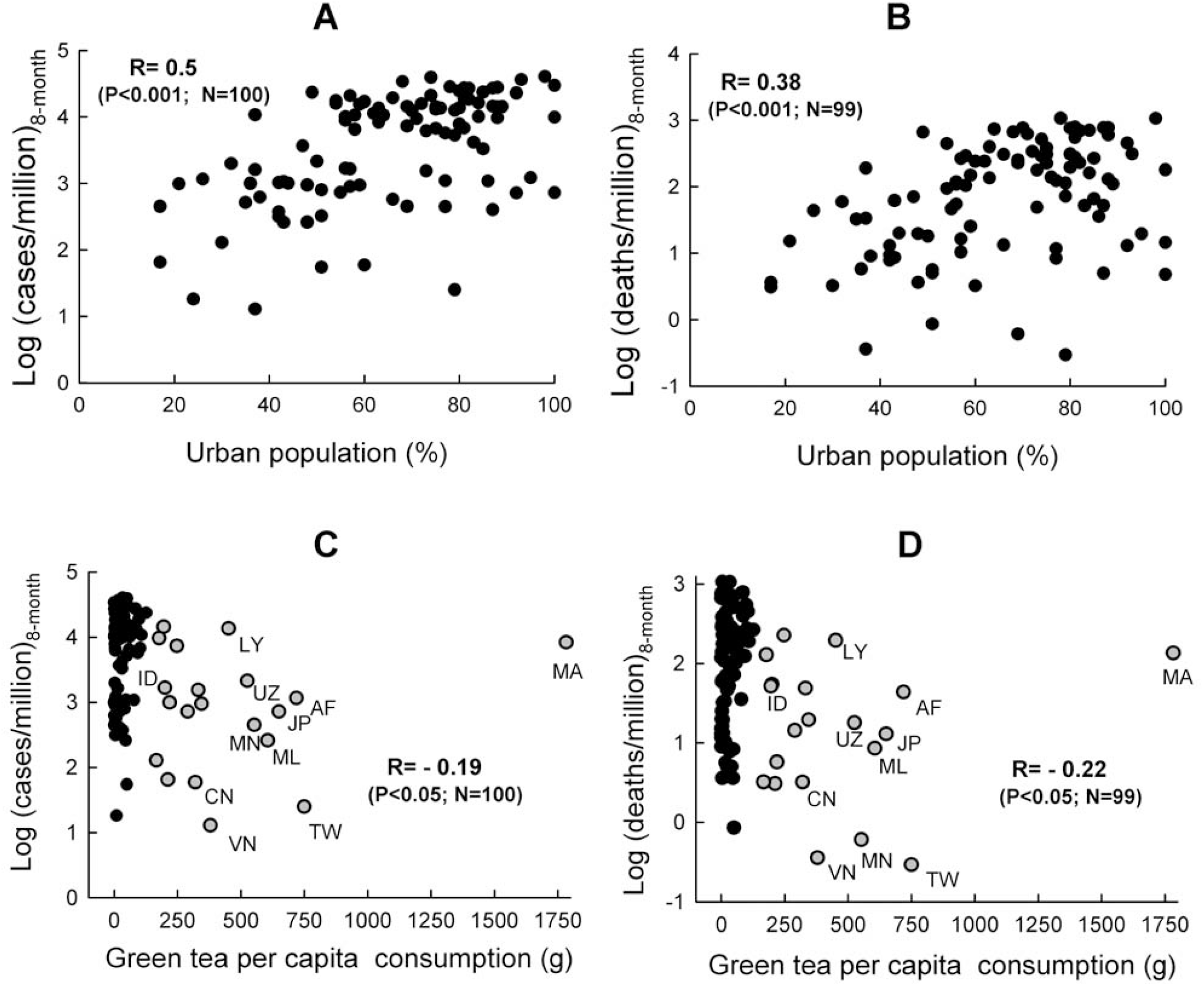
Percentage of urban population and per capita green tea consumption are associated with COVID-19 morbidity and mortality. COVID-19 morbidity (**A, C**) and mortality (**B, D**) in relation to % of urban population (upper panel) and green tea consumption (lower panel). Morbidity and mortality values are adjusted for the onset of the disease (8 months after the onset). Countries with values available as of January 22, 2021 were included. Values of regression coefficient R and significance levels denoted were calculated using the multiple linear regression model, N –number of countries included in the analysis. See text for more details. On plots **C** and **D** gray circles represent values for the countries with ‘high’ green tea consumption; for some of these standard two letter country codes are provided. AF- Afghanistan; CN –China; ID – Indonesia; JP –Japan; LY- Libya; MA- Morocco; ML –Mali; MN-Mongolia; TW- Taiwan; UZ – Uzbekistan. Notice, the circle representing Indonesia in **D**, is almost completely overlapped with that representing UAE.

Thus, it appears that COVID-19 morbidity and mortality are indeed associated with green tea consumption at the level of individual countries. Of note, these associations were still statistically significant when additional important [32] variable, namely Human Developmental Index (HDI) based on access to health and education services and income [31] was added to the model (see Supplementary **Table S3 (A**,**B)**).

It has been found recently that countries with routine mass drug administration of anti-parasitic prophylactic ivermectin have a significantly lower incidence of COVID-19 [34]. This important finding is in accordance with anti-viral activity of ivermectin in vitro [35] and other related evidence [36]. Therefore, mass drug administration of ivermectin could affect associations considered in the present study as a confounder. To account for this possibility, countries with mass drug administration of ivermectin [37] were excluded from the linear regression analysis and corresponding values were recalculated. In this restricted subset of countries, the associations between COVID-19 morbidity (and mortality) and green tea consumption were still significant (Supplementary **Table S4 (A, B)**. Moreover, the strengths of both correlations became notably higher (−0.24 and −0.28) as compared to the unrestricted set of countries (−0.19 and −0.22). These results indicate that the associations between COVID-19 morbidity (and mortality) and green tea consumption are not due to an overlap of countries with higher green tea consumption with countries with mass drug administration of ivermectin. Additionally, these results indirectly support previously reported association between mass drug administration of ivermectin and lower incidence of COVID-19 [34].

## Discussion

*The main finding* of this study is that higher green tea consumption is associated with lower COVID-19 morbidity/mortality at the level of ecological study. Indeed, there are striking differences in COVID-19 morbidity/mortality between groups of countries with ‘high’ and ‘low’ green tea consumption. The differences were still observed after the adjustment for the onset of the disease. Besides, the analysis using multiple linear regression approach suggests that the associations are present at the level of individual countries. At this level, this is presumably the first evidence supporting the idea that green tea constituents may reduce the risks related to COVID-19. These results are in agreement with several lines of emerging evidence implying that green tea catechins may be effective in prevention/treatment of COVID-19 or amelioration of its severity ([7-11,14]) (as outlined in the Introduction). Nevertheless, in the context of this study, it would be fair to mention that green catechins are not *the only* constituents of green tea. Other constituents may also be of importance for potential beneficial effects.

### Limitations of the study and potential future directions

For most of the countries green tea consumption was estimated based on the annual balance between its import and export. Although this approach is used for coarse estimates [22], apparently, it has several limitations as outlined in Methods. Thus, the list of countries with ‘high’ green tea consumption should be considered as a tentative. It cannot be excluded that some countries with considerable green tea per/capita consumption have not been identified among those producing green tea (for instance, Myanmar, South Korea, Papua New Guinea). It would be of interest to obtain relevant information in regard of these countries, include it in the analysis and check how this would affect associations considered in this work.

Other limitations include multiple factors that may differentially affect COVID-19 morbidity and mortality in distinct countries. For instance: the administrative strategies to prevent transmission; population density, percentage of urban population, percentage of older population; condition-specific mortality risks; TB infection and BCG vaccination [25,27, 38]. Potential role of only some of these factors was assessed in this study. On the other hand, potential confounding factors included in the linear regression model in this study represent the majority of these identified as *consistent* (and at the same time relatively independent) in relation to COVID-19 in previous studies [25, 32]. Additionally, it appears that there are *causal* relations between green tea consumption, and several condition-specific risks (such as risks of cardiovascular diseases [13], diabetes [11], etc). Therefore, inclusion of these risk factors in the regression model *as confounders* would be rather controversial because a confounding variable should be an independent factor for the outcome of interest [39]. It is important to notice that results of any ecological study, taken alone, could only provide the information about a correlation rather than a causal relation. Thus, experimental studies are required to either confirm or reject the potential causal relation. Importantly, green tea catechins are considered as safe by FDA and have been already investigated in experimental studies in relation to some infections. Moreover, results of at least one *randomized placebo-controlled* study [40] strongly suggest that green tea catechins can protect against acute upper respiratory tract infections thus, quite likely, infections induced by members of coronavirus family. Therefore, it may be expected that green tea catechins can also protect against the infection induced by SARS-CoV-2. If this is the case, it would have important implications. Possible efficacy of green tea catechins in treatment of COVID-19 and amelioration of its severity should be also assessed in observational and experimental epidemiological studies as potentially even more important.

## Conclusion

Evidence supporting the idea that green tea constituents could reduce overall risks related to COVID-19 has been obtained. The results are promising and are in line with emerging evidence from other studies including pharmacological ones. Nevertheless, because of limitations of this study the idea still should be considered as a hypothesis requiring further assessment. Several vaccines are currently validated for COVID-19 prevention and mass vaccination has already been started in many countries. Nevertheless, it is likely that development of efficient drug therapy that reduces COVID-19 severity/mortality would be important for rather prolonged time. In this context, the results obtained in this study may have significant implications.

## Data Availability

Data derived from public domain resources

## Declarations section

### Ethical Approval and Consent to participate

Not applicable

### Consent for publication

Not applicable

### Availability of supporting data

Data (mostly) available within the article or its supplementary materials. Additional data available on request from the author.

## Acknowledgements

I would like express my gratitude to personnel of the open resources for creating excellent opportunities for research.

## Supplementary materials

**Table S1.**
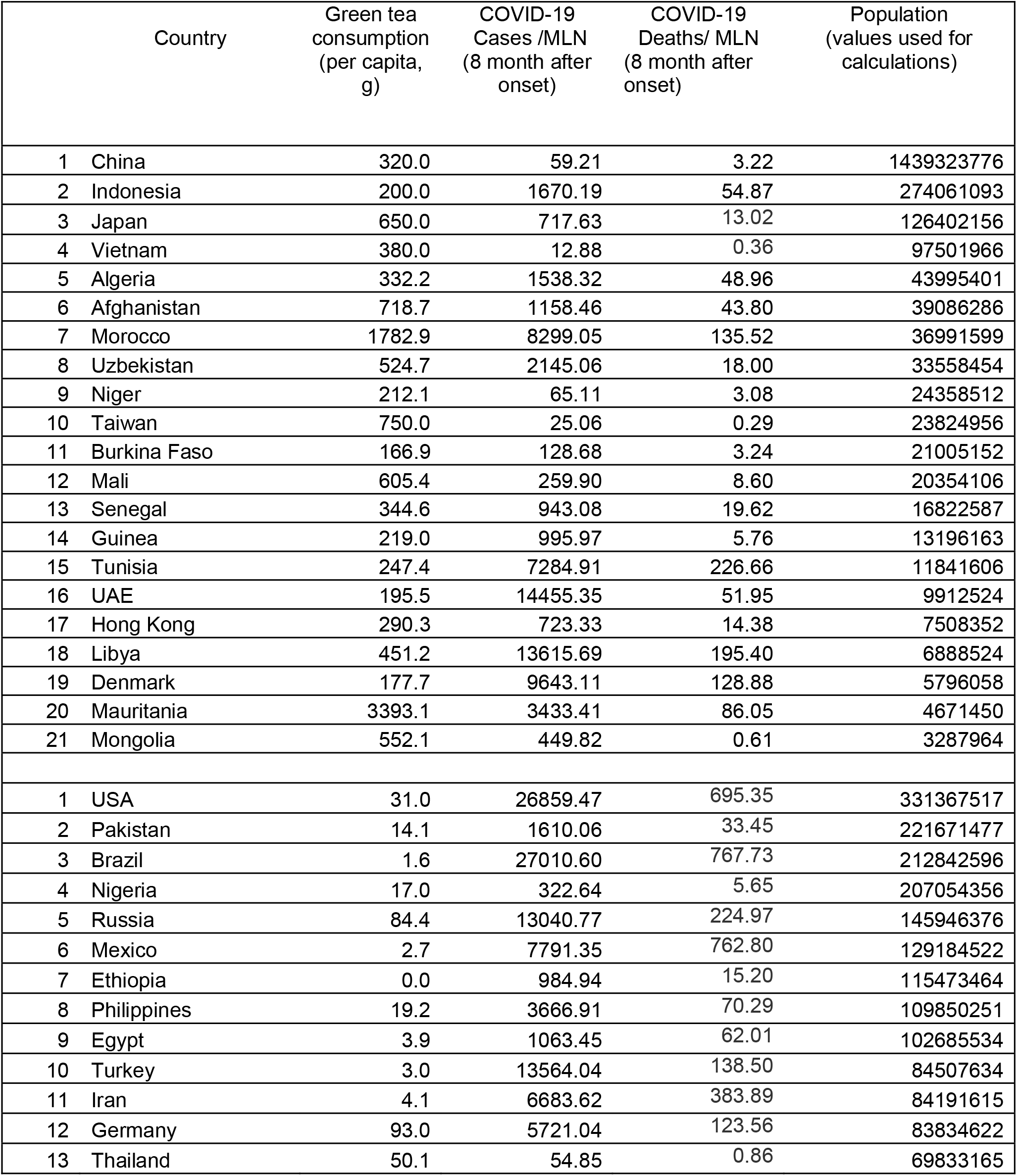
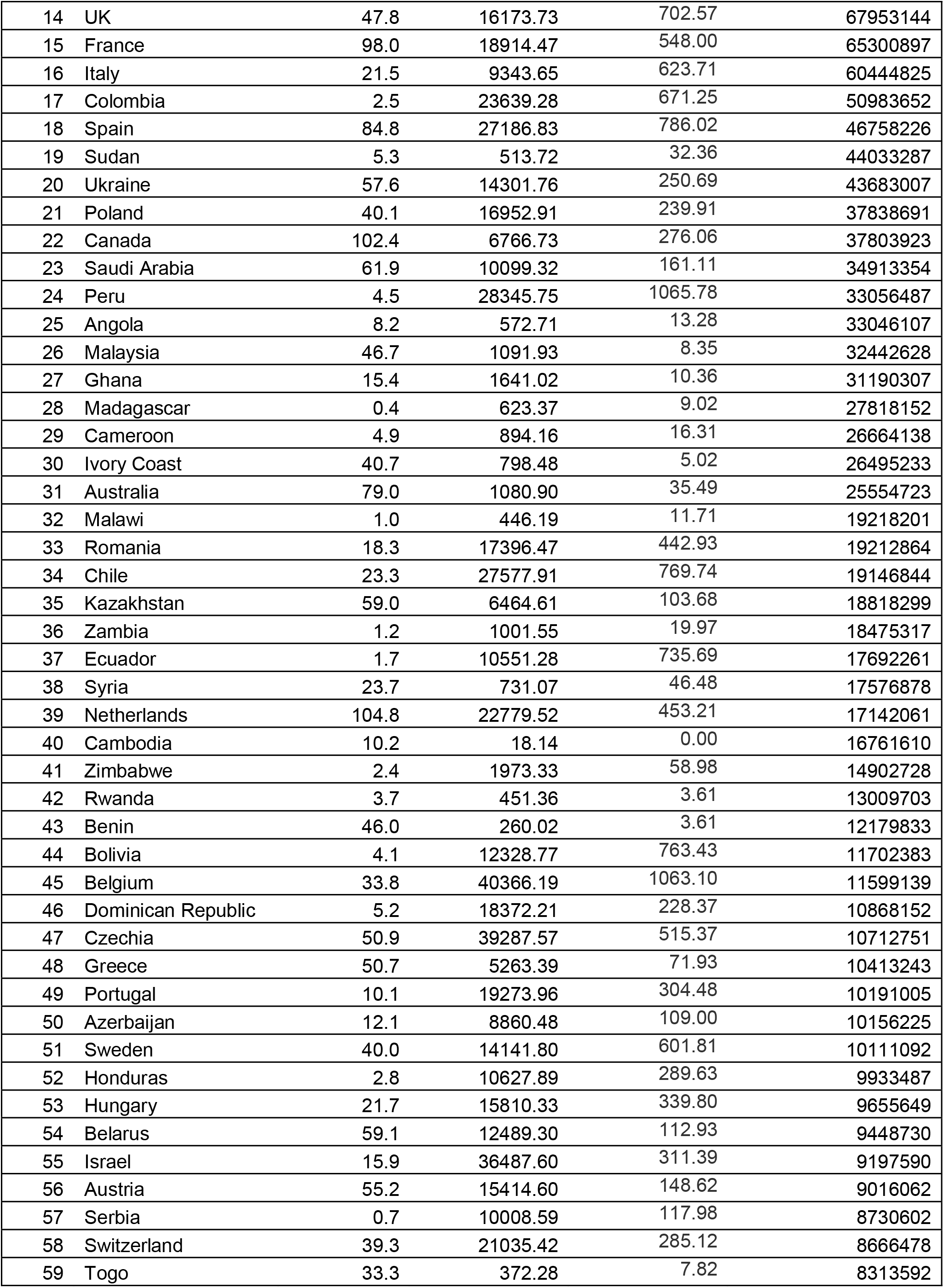
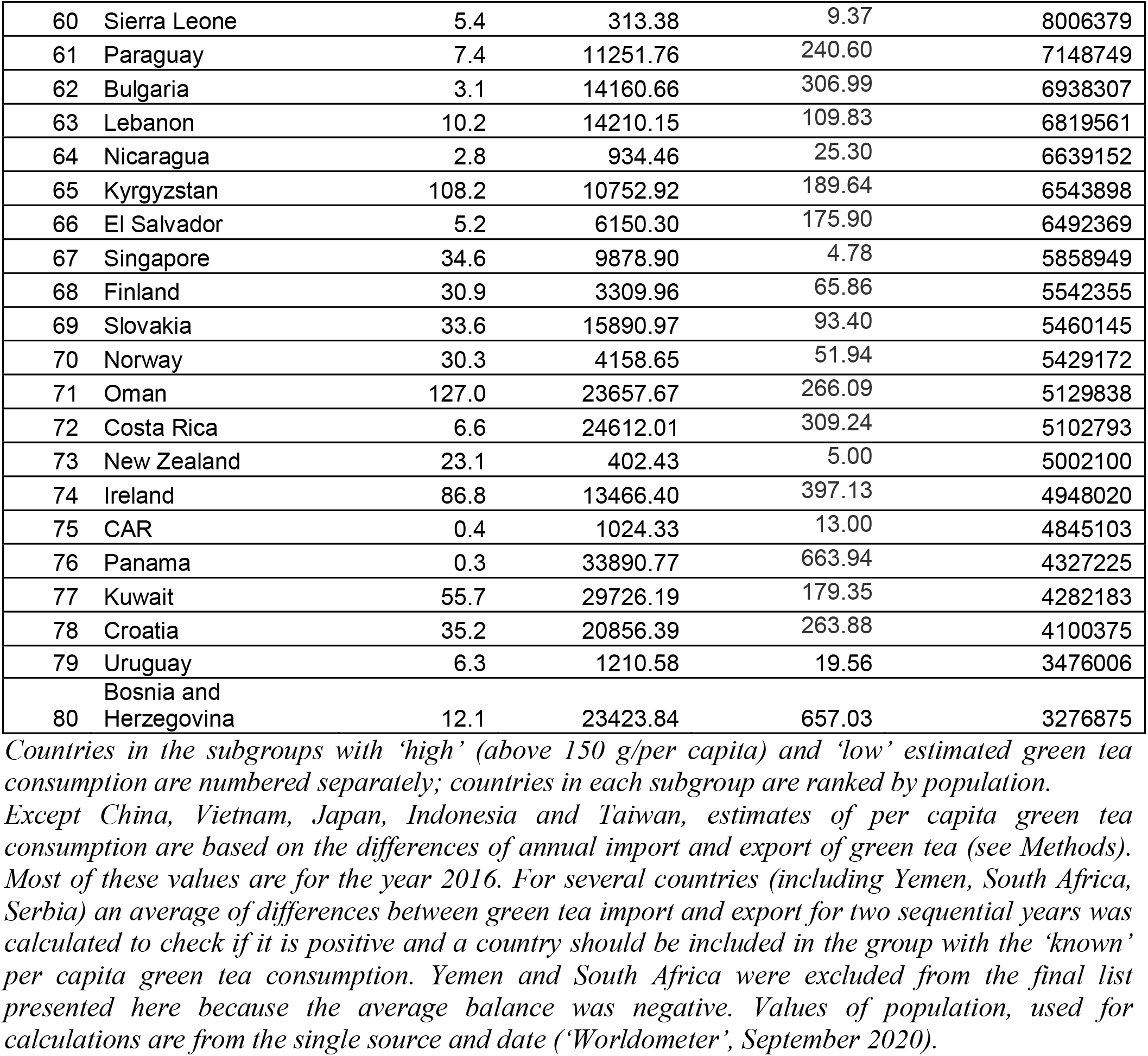
Countries included in the analysis of correlation using Spearman Rank Correlation.

| Country | Green tea consumption (per capita, g) | COVID-19 Cases /MLN (8 month after onset) | COVID-19 Deaths/ MLN (8 month after onset) | Population (values used for calculations) |
| --- | --- | --- | --- | --- |
| 1 China | 320.0 | 59.21 | 3.22 | 1439323776 |
| 2 Indonesia | 200.0 | 1670.19 | 54.87 | 274061093 |
| 3 Japan | 650.0 | 717.63 | 13.02 | 126402156 |
| 4 Vietnam | 380.0 | 12.88 | 0.36 | 97501966 |
| 5 Algeria | 332.2 | 1538.32 | 48.96 | 43995401 |
| 6 Afghanistan | 718.7 | 1158.46 | 43.80 | 39086286 |
| 7 Morocco | 1782.9 | 8299.05 | 135.52 | 36991599 |
| 8 Uzbekistan | 524.7 | 2145.06 | 18.00 | 33558454 |
| 9 Niger | 212.1 | 65.11 | 3.08 | 24358512 |
| 10 Taiwan | 750.0 | 25.06 | 0.29 | 23824956 |
| 11 Burkina Faso | 166.9 | 128.68 | 3.24 | 21005152 |
| 12 Mali | 605.4 | 259.90 | 8.60 | 20354106 |
| 13 Senegal | 344.6 | 943.08 | 19.62 | 16822587 |
| 14 Guinea | 219.0 | 995.97 | 5.76 | 13196163 |
| 15 Tunisia | 247.4 | 7284.91 | 226.66 | 11841606 |
| 16 UAE | 195.5 | 14455.35 | 51.95 | 9912524 |
| 17 Hong Kong | 290.3 | 723.33 | 14.38 | 7508352 |
| 18 Libya | 451.2 | 13615.69 | 195.40 | 6888524 |
| 19 Denmark | 177.7 | 9643.11 | 128.88 | 5796058 |
| 20 Mauritania | 3393.1 | 3433.41 | 86.05 | 4671450 |
| 21 Mongolia | 552.1 | 449.82 | 0.61 | 3287964 |
| 1 USA | 31.0 | 26859.47 | 695.35 | 331367517 |
| 2 Pakistan | 14.1 | 1610.06 | 33.45 | 221671477 |
| 3 Brazil | 1.6 | 27010.60 | 767.73 | 212842596 |
| 4 Nigeria | 17.0 | 322.64 | 5.65 | 207054356 |
| 5 Russia | 84.4 | 13040.77 | 224.97 | 145946376 |
| 6 Mexico | 2.7 | 7791.35 | 762.80 | 129184522 |
| 7 Ethiopia | 0.0 | 984.94 | 15.20 | 115473464 |
| 8 Philippines | 19.2 | 3666.91 | 70.29 | 109850251 |
| 9 Egypt | 3.9 | 1063.45 | 62.01 | 102685534 |
| 10 Turkey | 3.0 | 13564.04 | 138.50 | 84507634 |
| 11 Iran | 4.1 | 6683.62 | 383.89 | 84191615 |
| 12 Germany | 93.0 | 5721.04 | 123.56 | 83834622 |
| 13 Thailand | 50.1 | 54.85 | 0.86 | 69833165 |

|  |  |  |  |  |  |
| --- | --- | --- | --- | --- | --- |
| 14 | UK | 47.8 | 16173.73 | 702.57 | 67953144 |
| 15 | France | 98.0 | 18914.47 | 548.00 | 65300897 |
| 16 | Italy | 21.5 | 9343.65 | 623.71 | 60444825 |
| 17 | Colombia | 2.5 | 23639.28 | 671.25 | 50983652 |
| 18 | Spain | 84.8 | 27186.83 | 786.02 | 46758226 |
| 19 | Sudan | 5.3 | 513.72 | 32.36 | 44033287 |
| 20 | Ukraine | 57.6 | 14301.76 | 250.69 | 43683007 |
| 21 | Poland | 40.1 | 16952.91 | 239.91 | 37838691 |
| 22 | Canada | 102.4 | 6766.73 | 276.06 | 37803923 |
| 23 | Saudi Arabia | 61.9 | 10099.32 | 161.11 | 34913354 |
| 24 | Peru | 4.5 | 28345.75 | 1065.78 | 33056487 |
| 25 | Angola | 8.2 | 572.71 | 13.28 | 33046107 |
| 26 | Malaysia | 46.7 | 1091.93 | 8.35 | 32442628 |
| 27 | Ghana | 15.4 | 1641.02 | 10.36 | 31190307 |
| 28 | Madagascar | 0.4 | 623.37 | 9.02 | 27818152 |
| 29 | Cameroon | 4.9 | 894.16 | 16.31 | 26664138 |
| 30 | Ivory Coast | 40.7 | 798.48 | 5.02 | 26495233 |
| 31 | Australia | 79.0 | 1080.90 | 35.49 | 25554723 |
| 32 | Malawi | 1.0 | 446.19 | 11.71 | 19218201 |
| 33 | Romania | 18.3 | 17396.47 | 442.93 | 19212864 |
| 34 | Chile | 23.3 | 27577.91 | 769.74 | 19146844 |
| 35 | Kazakhstan | 59.0 | 6464.61 | 103.68 | 18818299 |
| 36 | Zambia | 1.2 | 1001.55 | 19.97 | 18475317 |
| 37 | Ecuador | 1.7 | 10551.28 | 735.69 | 17692261 |
| 38 | Syria | 23.7 | 731.07 | 46.48 | 17576878 |
| 39 | Netherlands | 104.8 | 22779.52 | 453.21 | 17142061 |
| 40 | Cambodia | 10.2 | 18.14 | 0.00 | 16761610 |
| 41 | Zimbabwe | 2.4 | 1973.33 | 58.98 | 14902728 |
| 42 | Rwanda | 3.7 | 451.36 | 3.61 | 13009703 |
| 43 | Benin | 46.0 | 260.02 | 3.61 | 12179833 |
| 44 | Bolivia | 4.1 | 12328.77 | 763.43 | 11702383 |
| 45 | Belgium | 33.8 | 40366.19 | 1063.10 | 11599139 |
| 46 | Dominican Republic | 5.2 | 18372.21 | 228.37 | 10868152 |
| 47 | Czechia | 50.9 | 39287.57 | 515.37 | 10712751 |
| 48 | Greece | 50.7 | 5263.39 | 71.93 | 10413243 |
| 49 | Portugal | 10.1 | 19273.96 | 304.48 | 10191005 |
| 50 | Azerbaijan | 12.1 | 8860.48 | 109.00 | 10156225 |
| 51 | Sweden | 40.0 | 14141.80 | 601.81 | 10111092 |
| 52 | Honduras | 2.8 | 10627.89 | 289.63 | 9933487 |
| 53 | Hungary | 21.7 | 15810.33 | 339.80 | 9655649 |
| 54 | Belarus | 59.1 | 12489.30 | 112.93 | 9448730 |
| 55 | Israel | 15.9 | 36487.60 | 311.39 | 9197590 |
| 56 | Austria | 55.2 | 15414.60 | 148.62 | 9016062 |
| 57 | Serbia | 0.7 | 10008.59 | 117.98 | 8730602 |
| 58 | Switzerland | 39.3 | 21035.42 | 285.12 | 8666478 |
| 59 | Togo | 33.3 | 372.28 | 7.82 | 8313592 |
| 60 | Sierra Leone | 5.4 | 313.38 | 9.37 | 8006379 |
| 61 | Paraguay | 7.4 | 11251.76 | 240.60 | 7148749 |
| 62 | Bulgaria | 3.1 | 14160.66 | 306.99 | 6938307 |
| 63 | Lebanon | 10.2 | 14210.15 | 109.83 | 6819561 |
| 64 | Nicaragua | 2.8 | 934.46 | 25.30 | 6639152 |
| 65 | Kyrgyzstan | 108.2 | 10752.92 | 189.64 | 6543898 |
| 66 | El Salvador | 5.2 | 6150.30 | 175.90 | 6492369 |
| 67 | Singapore | 34.6 | 9878.90 | 4.78 | 5858949 |
| 68 | Finland | 30.9 | 3309.96 | 65.86 | 5542355 |
| 69 | Slovakia | 33.6 | 15890.97 | 93.40 | 5460145 |
| 70 | Norway | 30.3 | 4158.65 | 51.94 | 5429172 |
| 71 | Oman | 127.0 | 23657.67 | 266.09 | 5129838 |
| 72 | Costa Rica | 6.6 | 24612.01 | 309.24 | 5102793 |
| 73 | New Zealand | 23.1 | 402.43 | 5.00 | 5002100 |
| 74 | Ireland | 86.8 | 13466.40 | 397.13 | 4948020 |
| 75 | CAR | 0.4 | 1024.33 | 13.00 | 4845103 |
| 76 | Panama | 0.3 | 33890.77 | 663.94 | 4327225 |
| 77 | Kuwait | 55.7 | 29726.19 | 179.35 | 4282183 |
| 78 | Croatia | 35.2 | 20856.39 | 263.88 | 4100375 |
| 79 | Uruguay | 6.3 | 1210.58 | 19.56 | 3476006 |
| 80 | Bosnia and Herzegovina | 12.1 | 23423.84 | 657.03 | 3276875 |

*Countries in the subgroups with 'high' (above 150 g/per capita) and 'low' estimated green tea consumption are numbered separately; countries in each subgroup are ranked by population.*
*Except China, Vietnam, Japan, Indonesia and Taiwan, estimates of per capita green tea consumption are based on the differences of annual import and export of green tea (see Methods). Most of these values are for the year 2016. For several countries (including Yemen, South Africa, Serbia) an average of differences between green tea import and export for two sequential years was calculated to check if it is positive and a country should be included in the group with the 'known' per capita green tea consumption. Yemen and South Africa were excluded from the final list presented here because the average balance was negative. Values of population, used for calculations are from the single source and date ('Worldometer', September 2020).*

**Table S2.** A. Higher per capita green tea consumption is associated with lower COVID-19 morbidity.

| Regression Coefficient |  |  |  |  |  |  |  |  |
| --- | --- | --- | --- | --- | --- | --- | --- | --- |
|  | Estimate | SE | t(cal) |  | P(T<=t(cal)) | Lower 95% | Upper 95% | VIF (Variance Inflation Factor) |
| b0 | 2.139633 | 0.234946 | 9.1069164 | *** (P<=0.001) | 1.34E-14 | 1.673206 | 2.60606 |  |
| b1:X1 | -0.00067 | 0.000287 | -2.321013 | * (P<=0.05) | 0.02242481 | -0.00124 | -9.65E-05 | 1.00669 |
| b2:X2 | -0.00012 | 6.66E-05 | -1.877445 | N.S. (P>0.05) | 0.06352517 | -0.00026 | 7.18E-06 | 1.075085 |
| b3:X3 | 0.017188 | 0.011256 | 1.5270234 | N.S. (P>0.05) | 0.13007786 | -0.00516 | 0.039534 | 1.364101 |
| b4:X4 | 0.02038 | 0.003988 | 5.1105683 | *** (P<=0.001) | 1.66E-06 | 0.012463 | 0.028296 | 1.443678 |
| Standard Regression Coefficient |  |  |  |  |  |  |  |  |
|  | Estimate | SE |  |  |  |  |  |  |
| b1:X1 | -0.18913 | 0.081484 |  |  |  | X1 | green tea consumption |  |
| b2:X2 | -0.15809 | 0.084207 |  |  |  | X2 | population density |  |
| b3:X3 | 0.144842 | 0.094852 |  |  |  | X3 | %population aged above 65 |  |
| b4:X4 | 0.498688 | 0.09758 |  |  |  | X4 | urban population % of total |  |
|  |  |  |  |  |  | Y | Log(cases/million)<sub>8-month</sub> |  |

**B. Higher per capita green tea consumption is associated with lower COVID-19 mortality.**
| Regression Coefficient |  |  |  |  |  |  |  |  |
| --- | --- | --- | --- | --- | --- | --- | --- | --- |
|  | Estimate | SE | t(cal) |  | P(T<=t(cal)) | Lower 95% | Upper 95% | VIF (Variance Inflation Factor) |
| b0 | 0.656052 | 0.251973 | 2.6036558 | * (P<=0.05) | 0.01071966 | 0.155753 | 1.156351 |  |
| b1:X1 | -0.00078 | 0.0003 | -2.609203 | * (P<=0.05) | 0.0105592 | -0.00138 | -0.00019 | 1.007759 |
| b2:X2 | -0.00023 | 6.94E-05 | -3.289652 | ** (P<=0.01) | 0.00141276 | -0.00037 | -9.06E-05 | 1.076687 |
| b3:X3 | 0.025541 | 0.011739 | 2.1757647 | * (P<=0.05) | 0.03208023 | 0.002233 | 0.048849 | 1.35754 |
| b4:X4 | 0.016141 | 0.004241 | 3.805876 | *** (P<=0.001) | 0.0002515 | 0.00772 | 0.024561 | 1.439814 |
| Standard Regression Coefficient |  |  |  |  |  |  |  |  |
|  | Estimate | SE |  |  |  |  |  |  |
| b1:X1 | -0.21781 | 0.083476 |  |  |  | X1 | green tea consumption |  |
| b2:X2 | -0.28384 | 0.086284 |  |  |  | X2 | population density |  |
| b3:X3 | 0.210801 | 0.096886 |  |  |  | X3 | %population aged above 65 |  |
| b4:X4 | 0.379746 | 0.099779 |  |  |  | X4 | urban population % of total |  |
|  |  |  |  |  |  | Y | Log(deaths/million)<sub>8-month</sub> |  |

In these and the following Tables the values are copy-pasted from 'KyPlot' (used for multiple regression analysis).

**Table S3.**

|  | Estimate | SE | t(cal) |  | P(T<=t(cal)) | Lower 95% | Upper 95% | VIF (Variance Inflation Factor) |
| --- | --- | --- | --- | --- | --- | --- | --- | --- |
| b0 | 1.299701 | 0.410205 | 3.1684225 | ** (P<=0.01) | 0.00206818 | 0.485231 | 2.114172 |  |
| b1:X1 | -0.00066 | 0.00028 | -2.352308 | * (P<=0.05) | 0.02074334 | -0.00121 | -1.03E-04 | 1.006838 |
| b2:X2 | -0.00013 | 6.49E-05 | -1.993778 | * (P<=0.05) | 0.04907418 | -0.00026 | -5.36E-07 | 1.075888 |
| b3:X3 | -0.00863 | 0.015156 | -0.569096 | N.S. (P>0.05) | 0.5706482 | -0.03872 | 0.021467 | 2.60554 |
| b4:X4 | 0.011593 | 0.00527 | 2.1997605 | * (P<=0.05) | 0.0302754 | 0.001129 | 0.022057 | 2.656559 |
| b5:X5 | 2.262758 | 0.91704 | 2.4674591 | * (P<=0.05) | 0.01541704 | 0.441954 | 4.083561 | 4.741674 |

Standard Regression Coefficient
|  | Estimate | SE |  |  |
| --- | --- | --- | --- | --- |
| b1:X1 | -0.18675 | 0.079392 | X1 | green tea consumption |
| b2:X2 | -0.16363 | 0.082069 | X2 | population density |
| b3:X3 | -0.07268 | 0.127715 | X3 | %population aged above 65 |
| b4:X4 | 0.283681 | 0.12896 | X4 | urban population % of total |
| b5:X5 | 0.425119 | 0.17229 | X5 | HDI |
|  |  |  | Y | Log(cases/million)<sub>8-month</sub> |

|  | Estimate | SE | t(cal) |  | P(T<=t(cal)) | Lower 95% | Upper 95% | VIF (Variance Inflation Factor) |
| --- | --- | --- | --- | --- | --- | --- | --- | --- |
| b0 | 0.054116 | 0.435535 | 0.1242522 | N.S. (P>0.05) | 0.90138373 | -0.81077 | 0.919002 |  |
| b1:X1 | -0.00078 | 0.000297 | -2.62E+00 | * (P<=0.05) | 0.01020825 | -0.00137 | -0.00019 | 1.007814 |
| b2:X2 | -0.00023 | 6.88E-05 | -3.360932 | ** (P<=0.01) | 0.00112832 | -0.00037 | -9.46E-05 | 1.077296 |
| b3:X3 | 0.006836 | 0.016069 | 0.4254099 | N.S. (P>0.05) | 0.67152071 | -0.02507 | 0.038745 | 2.593504 |
| b4:X4 | 0.009634 | 0.005703 | 1.6890875 | N.S. (P>0.05) | 0.09455285 | -0.00169 | 0.02096 | 2.655167 |
| b5:X5 | 1.644549 | 0.975253 | 1.6862791 | N.S. (P>0.05) | 0.09509418 | -0.29211 | 3.581209 | 4.72475 |

Standard Regression Coefficient
|  | Estimate | SE |  |  |
| --- | --- | --- | --- | --- |
| b1:X1 | -0.21678 | 0.082672 | X1 | green tea consumption |
| b2:X2 | -0.28727 | 0.085474 | X2 | population density |
| b3:X3 | 0.056418 | 0.132621 | X3 | %population aged above 65 |
| b4:X4 | 0.226655 | 0.134188 | X4 | urban population % of total |
| b5:X5 | 0.301847 | 0.179002 | X5 | HDI |
|  |  |  | Y | Log(deaths/million)<sub>8-month</sub> |

**Table S4.**

|  | Estimate | SE | t(cal) |  | P(T<=t(cal)) | Lower<br>95% | Upper<br>95% | VIF (Variance<br>Inflation Factor) |
| --- | --- | --- | --- | --- | --- | --- | --- | --- |
| b0 | 2.416337 | 0.28836 | 8.3795856 | *** (P<=0.001) | 1.34E-12 | 1.842591 | 2.990083 |  |
| b1:X1 | -0.00077 | 0.000301 | -2.543183 | * (P<=0.05) | 0.01288754 | -0.00137 | -0.00017 | 1.015859 |
| b2:X2 | -0.00012 | 6.87E-05 | -1.799757 | N.S. (P>0.05) | 0.07562258 | -0.00026 | 1.30E-05 | 1.078535 |
| b3:X3 | 0.00807 | 0.01215 | 0.6642211 | N.S. (P>0.05) | 0.50843524 | -0.0161 | 0.032245 | 1.251498 |
| b4:X4 | 0.018918 | 0.004494 | 4.2099507 | *** (P<=0.001) | 6.57E-05 | 0.009977 | 0.027858 | 1.328393 |

Standard Regression Coefficient
|  | Estimate | SE |  |  |
| --- | --- | --- | --- | --- |
| b1:X1 | -0.23806 | 0.093607 | X1 | green tea consumption |
| b2:X2 | -0.17359 | 0.096451 | X2 | population density |
| b3:X3 | 0.069011 | 0.103898 | X3 | %population aged above 65 |
| b4:X4 | 0.450642 | 0.107042 | X4 | urban population % of total |
|  |  |  | Y | Log(cases/million)<sub>8-month</sub> |

|  | Estimate | SE | t(cal) |  | P(T<=t(cal)) | Lower<br>95% | Upper<br>95% | VIF (Variance<br>Inflation Factor) |
| --- | --- | --- | --- | --- | --- | --- | --- | --- |
| b0 | 0.995125 | 0.307901 | 3.2319609 | ** (P<=0.01) | 0.00178623 | 0.382382 | 1.607868 |  |
| b1:X1 | -0.00093 | 0.00031 | -2.994489 | ** (P<=0.01) | 0.00365727 | -0.00154 | -0.00031 | 1.0188 |
| b2:X2 | -0.00023 | 7.04E-05 | -3.225352 | ** (P<=0.01) | 0.00182307 | -0.00037 | -8.70E-05 | 1.081253 |
| b3:X3 | 0.013841 | 0.012442 | 1.1124154 | N.S. (P>0.05) | 0.26929123 | -0.01092 | 0.038601 | 1.238439 |
| b4:X4 | 0.014528 | 0.00474 | 3.0648789 | ** (P<=0.01) | 0.00296827 | 0.005095 | 0.02396 | 1.31987 |

Standard Regression Coefficient
|  | Estimate | SE |  |  |
| --- | --- | --- | --- | --- |
| b1:X1 | -0.28244 | 0.094319 | X1 | green tea consumption |
| b2:X2 | -0.3134 | 0.097167 | X2 | population density |
| b3:X3 | 0.11568 | 0.10399 | X3 | %population aged above 65 |
| b4:X4 | 0.329028 | 0.107354 | X4 | urban population % of total |
|  |  |  | Y | Log(deaths/million)<sub>8-month</sub> |

<sup>1</sup> Countries with mass drug administration of ivermectin: Angola, Benin, Burcina Faso, Cameron, Ivory Coast, CAR, Ethiopia, Ghana, Niger, Nigeria, Senegal, Sierra Leone, Sudan, Malaysia.

